# Prediction of Celiac Disease Severity and Associated Endocrine Morbidities through Deep Learning-based Image Analytics

**DOI:** 10.1101/2021.01.20.21250194

**Authors:** Lubaina Ehsan, Marium Khan, Rasoul Sali, Alexis M. Catalano, William Adorno, Kamran Kowsari, Lin Cheng, Patcharin Pramoonjago, Shyam Raghavan, Jocelyn Silvester, Mark DeBoer, Christopher A. Moskaluk, Sean R. Moore, Donald E. Brown, Sana Syed

**Affiliations:** Division of Gastroenterology, Hepatology and Nutrition, Department of Pediatrics, School of Medicine, University of Virginia, Charlottesville, VA; Department of Pediatrics, School of Medicine, University of Virginia, Charlottesville, VA; Department of System and Information Engineering, University of Virginia, Charlottesville, VA; University of Virginia, Charlottesville, VA; College of Dental Medicine, Columbia University, New York City, NY, USA; Pathology Department, Rush University Medical Center, Chicago, IL; Department of Pathology, School of Medicine, University of Virginia, Charlottesville, VA; Division of Gastroenterology, Hepatology and Nutrition, Boston Children’s Hospital, Boston, MA; Division of Endocrinology and Diabetes, Department of Pediatrics, School of Medicine, University of Virginia, Charlottesville, VA

## Abstract

**Objective:** Develop a deep learning-based methodology using the foundations of systems pathology to generate highly accurate predictive tools for complex gastrointestinal diseases, using celiac disease (CD) as a prototype.

**Design:** To predict the severity of CD, defined by Marsh–Oberhüber classification, we used deep learning to develop a model based on histopathologic features.

**Results:** The study was based on a pediatric cohort of 124 patients identified with different classes of CD severity. The model predicted CD with an overall 88.7% accuracy with the highest for Marsh IIIc (91.0%; 95% sensitivity; 91% specificity). The model identified EECs as a defining feature of children with Marsh IIIc CD and endocrinopathies which was confirmed using immunohistochemistry.

**Conclusion:** This deep learning image analysis platform has broad applications in disease treatment, management, and prognostication and paves the way for precision medicine.

**Summary:** *What is already known about this subject?:*

– Deep Learning has the potential to generate predictive models for complex gastrointestinal diseases.

*What are the new findings?:*

– Our deep learning-based model used the foundations of systems pathology to generate a highly accurate predictive tool for complex gastrointestinal diseases, using a celiac disease (CD) pediatric cohort as a prototype.
– The model predicated CD severity with high accuracy and identified enteroendocrine cells as a defining feature of children with severe CD and endocrinopathies.

*How might it impact on clinical practice in the foreseeable future?:*

– Assessment of histopathological markers at the time of diagnosis that can predict risk of severity or complications can have broad applications in disease treatment, management, and prognostication and pave the way for precision medicine.

## Introduction

Several Celiac Disease (CD) histopathological features and serologic tests the diagnose disease. Biopsy-based assessment of proximal small intestine is the current is the gold standard for confirmation of CD [1, 2, 3], with severity assessment using Marsh–Oberhüber classification (modified Marsh score) being common in clinical practice [4, 5]. Serologic tests are frequently used to identify individuals who require a biopsy; these include anti-gliadin IgA and IgG (AGA IgA and AGA IgG), anti-reticulin IgA (ARA), anti-endomysium IgA(EMA), and anti-tissue transglutaminase IgA (TTG) antibodies [1]. Despite there being guidelines for CD diagnosis, its severity assessment is challenging. It falls privy to interobserver variability, non-specificity of discerning histopathological features, and a lack of specific histopathological markers associated with either CD severity classes or comorbid conditions.

The Marsh–Oberhüber classification is a commonly used method for CD severity assessment in clinical practice based on histopathological features involving villus and crypt injury and intraepithelial inflammation [4, 5]. Unfortunately, high interobserver variability is found when this system is applied to clinical samples, and agreement is generally only fair (κ = 0.29-0.35) [6, 7].

Alternate purely quantitative methods of assessing mucosal damage by direct measurement of the villous height to crypt depth ratio (Vh: Cd) and intraepithelial lymphocyte (IEL) counts have been proposed [8]. Using these methods, the inter-observer agreement is moderate (κ = 0.55) [7]; however, this method is also highly dependent on the use of correctly oriented specimens and is not used in clinical practice.

In patients with CD, there is also evidence of comorbid endocrinopathies such as T1DM and hypothyroidism, which substantially more common than in the general population. Further, in comparison to those with isolated T1DM, patients with undiagnosed CD and T1DM also have a higher prevalence of retinopathy (58% vs. 25%) and nephropathy (42% vs. 4%) [9, 10, 11].

We chose a deep learning-based methodology for image analytics to develop a novel predictive model for CD severity and the association of disease features with comorbid conditions. These models utilizing Convolutional Neural Networks (CNNs) have shown potential benefit for disease characterization using histological specimens in a broad range of diseases [12, 13, 14, 15], including CD [16]. They are optimized to discern fine features from regions of interest within the biopsy images. One such example is of the deep residual network (ResNet) that has outperformed early deep learning models and achieved superior performance on image recognition benchmarks [17, 18, 19]. We also deployed a deep learning method [20] to provide insight into the model decision-making and discerning specific histologic features for each class. We confirmed the features identified by the model, specific for severe CD (Marsh IIIc) and concurrent endocrine commodities, using immunohistochemistry, which confirmed the utility of deep learning CNN models for discerning disease-specific features.

## Methods

### Participants and Data Collection

Duodenal biopsy glass slides from children (under the age of 18 years) at the time of CD diagnosis (baseline) from 1992 to 2017 (25 years) were retrieved at the University of Virginia (UVA), Charlottesville, Virginia. These were already Hematoxylin and eosin (H&E) stained as per standard pathological workflow. To digitize the glass biopsy slides, scanning was done at 40x magnification at the Biorepository and Tissue Research Facility, UVA, using a Leica SCN400 brightfield scanning instrument (Leica Microsystems CMS GmbH, Germany).

Biopsies selected and retrieved as CD were based on pathology reports. Histologically normal controls (referred to as controls for the purpose of this study) were based on the inclusion criteria of no evidence of disease (eosinophilic esophagitis, gastritis, inflammatory bowel disease, etc.) being present in any other part of the gastrointestinal tract examined via the esophagogastroduodenoscopy.

### Evaluation of Celiac Disease Severity

CD severity for each patient was assessed using the Marsh–Oberhüber classification, which involves evaluating duodenal villous architecture and intraepithelial lymphocytosis (Figure 1). In the case where more than one biopsy fragment was present per image, each fragment was separately assessed via the Marsh–Oberhüber classification. Details of the Marsh–Oberhüber classification (referred to as Marsh score as part of this study) are described in supplemental appendix **S1**.

**Figure 1:**
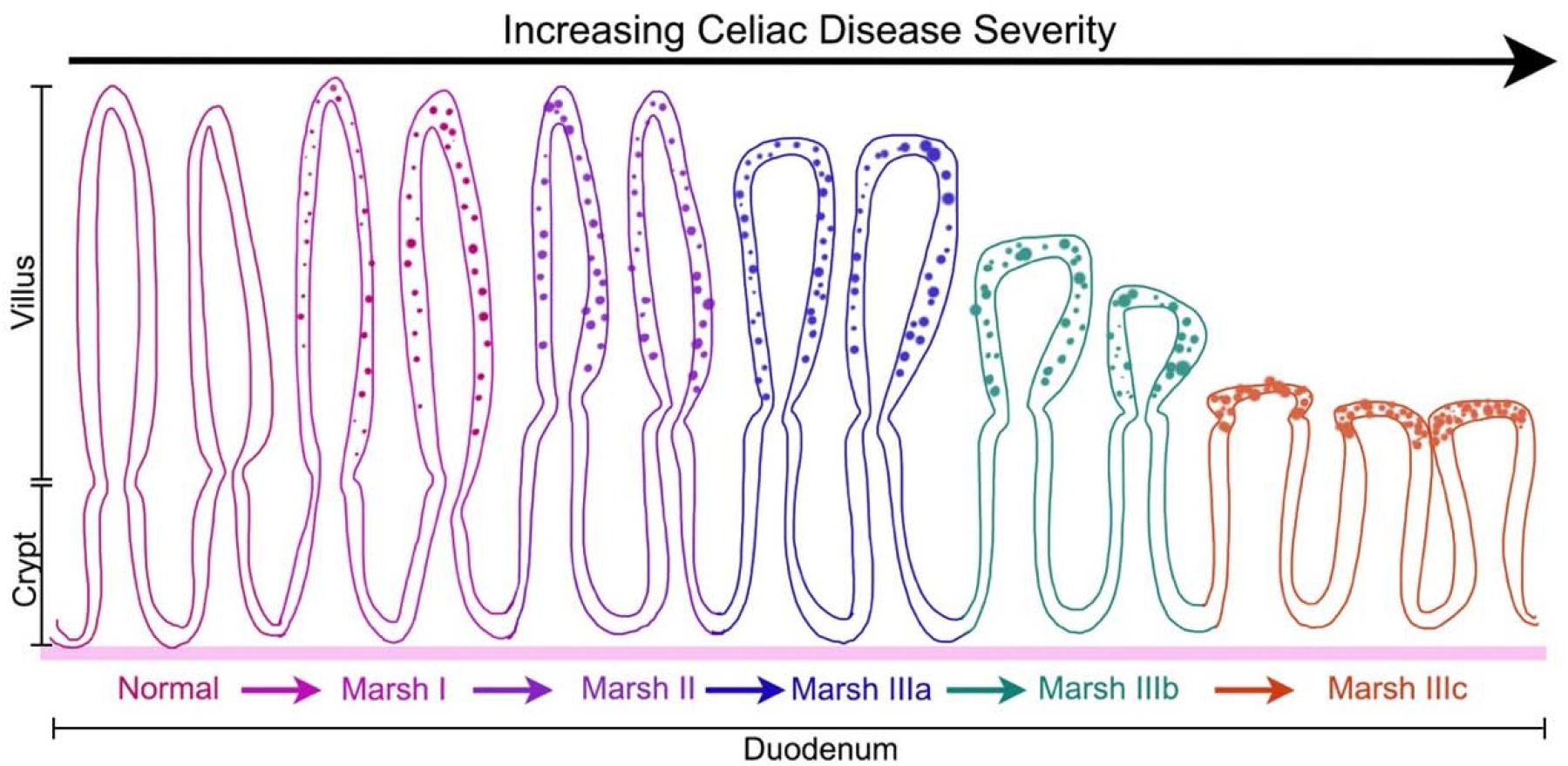
Illustrative explanation of the Marsh–Oberhüber classification for the duodenum.

### Deep Learning Image Analysis Model

#### Biopsy Image Model Input Dataset Construction

##### A. Biopsy Image Patch Creation

Approximately 50,000 patches were created using biopsy whole slide images (WSIs) to maximize of computational resources (details in supplemental appendix **S2**). Each patch was further rotated on its horizontal and vertical axes to augment the data sample.

##### B. Model Training and Test Set Development

Biopsy image patches were then randomly divided (70:30 ratio) into training and testing sets. Patient samples were kept separate among training and test sets without any overlap.

##### C. Biopsy Patch Clustering

A large proportion of the digital WSIs consist of white background, which does not provide clinically useful information and may bias the model. To overcome this, a two-step clustering process was applied to identify and remove patch clusters with more than 50% white background (figure and details in supplemental appendix **S3**).

##### D. Biopsy Stain Color Normalization

Slight variations in H&E staining chemical composition over the years result in visible H&E color differences. Therefore, stain color normalization as described by Vahadane et al. [21], was deployed to eliminate erroneous biopsy classification based on color differences rather than distinct biopsy image features. Details of this method are explained in supplemental appendix **S4**.

#### Deep Learning Model Development

First, a patch-level model was used to focus on microscopic histologic features in CD versus control biopsies. This was followed by a patch-level model for CD severity classes assessed via Marsh score only. Further, a WSI-level model was developed to obtain overall low magnification-level information, emulating the pathologist workflow of scanning zoomed out biopsy images to obtain an overall assessment. The detailed computational methods of our patch- and WSI-level models have been previously published [22].

##### A. Patch-level Model

ResNet50 trained on the ImageNet dataset was used. We customized Resnet50 by removing fully connected layers, retaining only the ResNet backbone as a feature extractor. Finally, a fully connected layer with 1024 neurons receiving a flattened output of feature extractor was added. The output layer represented the biopsy classification for each of the four Marsh score classes present in our data sample: I, IIIa, IIIb, and IIIc. Details of the model are described in the supplemental appendix **S5**.

##### B. Whole Slide Image-level Model

A heuristic approach was used for the WSI-level classification model. This approach aggregated patch-level classifications to provide WSI-level inferences. Computational details are described in the supplemental appendix **S6**.

#### Benchmark Visualization of Discerning Features for Model Classification

Features utilized by the image analysis model for decision making were visualized using Gradient-weighted Class Activation Mappings (Grad-CAMs). Grad-CAM computational details in supplemental appendix **S7**.

### Immunohistochemistry (IHC) Analysis

Biopsy slides were constructed from paraffin blocks. Immunohistochemistry was performed on a robotic platform (Ventana discover Ultra Staining Module, Ventana Co., Tucson, AZ, USA). A heat-induced antigen retrieval protocol set for 64 min was carried out using a TRIS– ethylenediamine tetraacetic acid (EDTA)–boric acid pH 8.4 buffer (Cell Conditioner 1). Endogenous peroxidases were blocked with peroxidase inhibitor (CM1) for 8 min before incubating the tissues with Chromogranin A antibody (Dako, Cat # A0430) at 1:400 dilution for 60 min at room temperature. The antigen-antibody complex was then detected using DISCOVERY anti-rabbit HQ HRP detection system and DISCOVERY ChromoMap DAB Kit (Ventana Co.). All the slides were counterstained with hematoxylin; they were dehydrated, cleared, and mounted for the assessment.

### IHC Qualitative and Quantitative Assessment

Pathologists confirmed qualitative assessment based on the color of the stained cells. Quantitative assessment was done using an unsupervised clustering algorithm called Gaussian Mixture Model (GMM), which groups biopsy image features using their pixel colors [23]. Our data was divided into five pixel color-based clusters, and details of the GMM method are mentioned in supplemental appendix **S8**.

### Descriptive Statistics

Biodemographic data were analyzed using IBM SPSS Statistics v. 22.0. Categorical variables were expressed as frequencies (%) and continuous as median with first and third quartile (Q1, Q3). Differences in the means of continuous variables were assessed via the Mann-Whitney U test and categorical variables using the Pearson Chi-Square test with p-value>0.05 as statistically significant. Anthropometric z-scores were calculated using reference guidelines and R application macros available through the World Health Organization.

### Ethical Considerations

This study was approved by the UVA Institutional Review Board (IRB #20448).

## Results

### Study Population for CD Severity Prediction

A total of 63 pediatric (under 18 years of age) CD patients (44.4% male) and 61 controls (54.1% male) with 239 and 174 whole-slide images (WSIs), respectively, were selected (3-4 biopsies images per patient). Population biodemographic characteristics are shown in **Table 1**. The biopsy image dataset encompassed disease severity ranging from Marsh I to IIIc (**Figure 1** shows an illustrative explanation of the Marsh–Oberhüber classification; also referred to as the modified Marsh score) with no biopsy images present with a modified Marsh score of II and it is known to be rare. The detailed distribution of the patch- and WSI-level biopsy image datasets are mentioned in supplemental appendix **S9**.

**Table 1:**
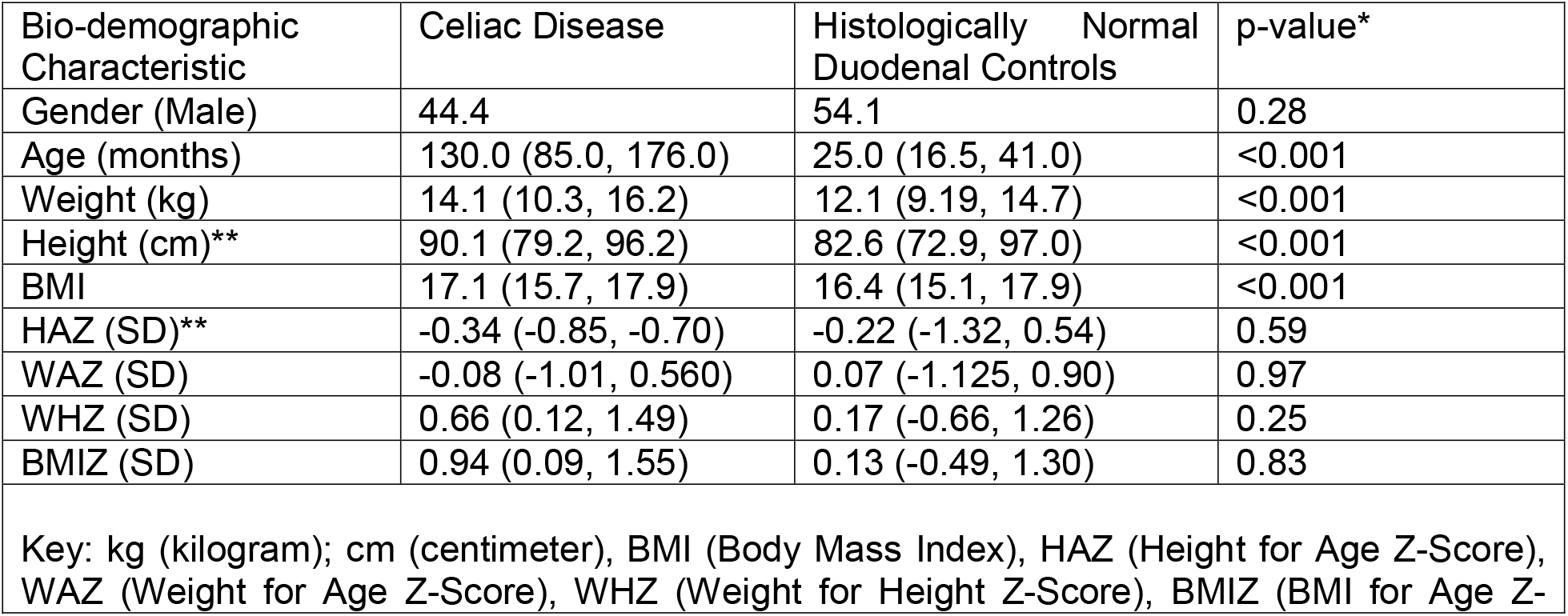
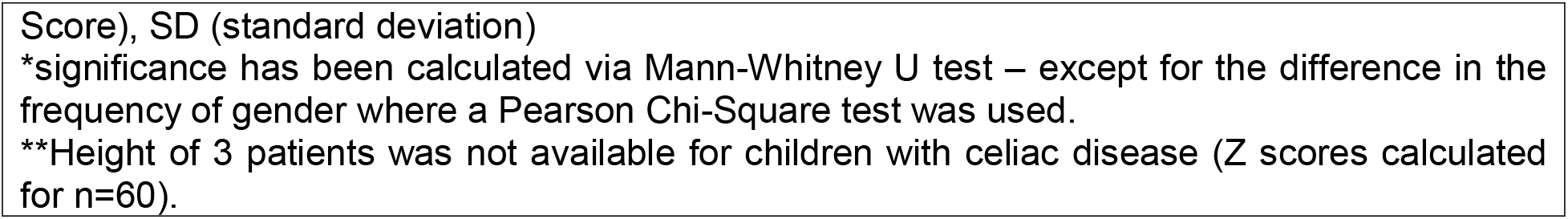
Pediatric Population Characteristics.

| Bio-demographic Characteristic | Celiac Disease | Histologically Normal Duodenal Controls | p-value* |
| --- | --- | --- | --- |
| Gender (Male) | 44.4 | 54.1 | 0.28 |
| Age (months) | 130.0 (85.0, 176.0) | 25.0 (16.5, 41.0) | <0.001 |
| Weight (kg) | 14.1 (10.3, 16.2) | 12.1 (9.19, 14.7) | <0.001 |
| Height (cm)** | 90.1 (79.2, 96.2) | 82.6 (72.9, 97.0) | <0.001 |
| BMI | 17.1 (15.7, 17.9) | 16.4 (15.1, 17.9) | <0.001 |
| HAZ (SD)** | -0.34 (-0.85, -0.70) | -0.22 (-1.32, 0.54) | 0.59 |
| WAZ (SD) | -0.08 (-1.01, 0.560) | 0.07 (-1.125, 0.90) | 0.97 |
| WHZ (SD) | 0.66 (0.12, 1.49) | 0.17 (-0.66, 1.26) | 0.25 |
| BMIZ (SD) | 0.94 (0.09, 1.55) | 0.13 (-0.49, 1.30) | 0.83 |

Key: kg (kilogram); cm (centimeter), BMI (Body Mass Index), HAZ (Height for Age Z-Score), WAZ (Weight for Age Z-Score), WHZ (Weight for Height Z-Score), BMIZ (BMI for Age Z-
Score), SD (standard deviation)
\*significance has been calculated via Mann-Whitney U test – except for the difference in the frequency of gender where a Pearson Chi-Square test was used.
\*\*Height of 3 patients was not available for children with celiac disease (Z scores calculated for n=60).

### Deep Learning Model Performance for CD Severity Prediction

The model for CD versus histologically normal controls demonstrated an accuracy of 94%, and the details are mentioned in supplemental appendix **S10**. For classifying CD Marsh-scored biopsy patches, the model had greater than 84% accuracy with the highest accuracy for Marsh IIIc (90.6%), as shown in **Figure 2** with performance metrics in **Table 2**. Receiver operating curves (ROC) and areas under the curve (AUC) for each Marsh score class are shown in **Figure 3**, where AUC for all classes was greater than 0.96. For WSIs, the model correctly classified each biopsy image with 100% accuracy via inference, as mentioned in the methods section.

**Table 2:** Performance metrics of the patch-level biopsy classification model.

| Class | Specificity | Sensitivity | PPV | NPV | Accuracy | F1 score | Recall | Precision |
| --- | --- | --- | --- | --- | --- | --- | --- | --- |
| Marsh I | 0.98 | 0.90 | 0.93 | 0.97 | 0.90 | 0.91 | 0.90 | 0.93 |
| Marsh IIIa | 0.98 | 0.85 | 0.94 | 0.96 | 0.85 | 0.89 | 0.85 | 0.94 |
| Marsh IIIb | 0.94 | 0.89 | 0.84 | 0.96 | 0.89 | 0.87 | 0.89 | 0.84 |
| Marsh IIIc | 0.95 | 0.91 | 0.86 | 0.97 | 0.91 | 0.88 | 0.91 | 0.86 |

Key: PPV (positive predictive value), NPV (negative predictive value)

**Figure 2:**
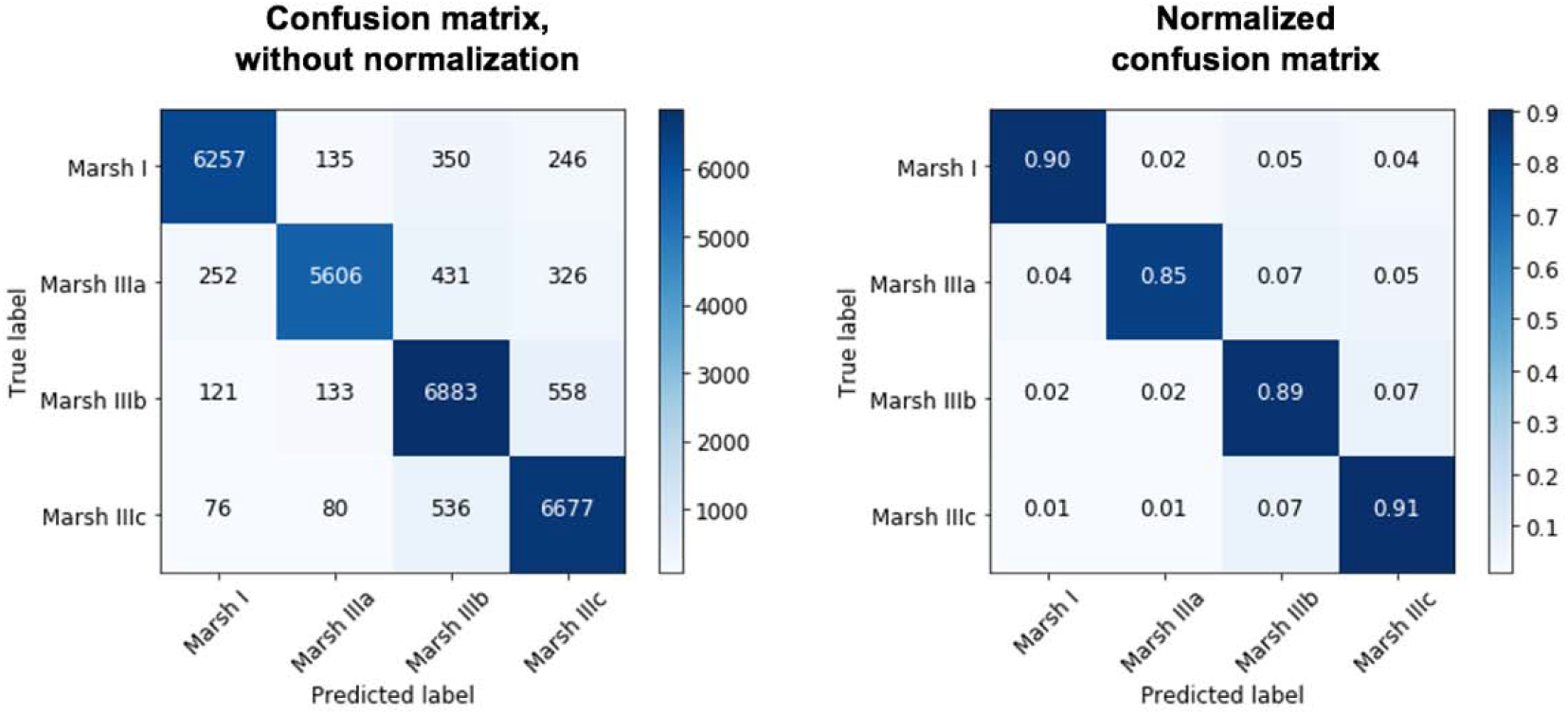
Confusion matrices for the patch-level performance of the model for celiac disease severity detection.

**Figure 3:**
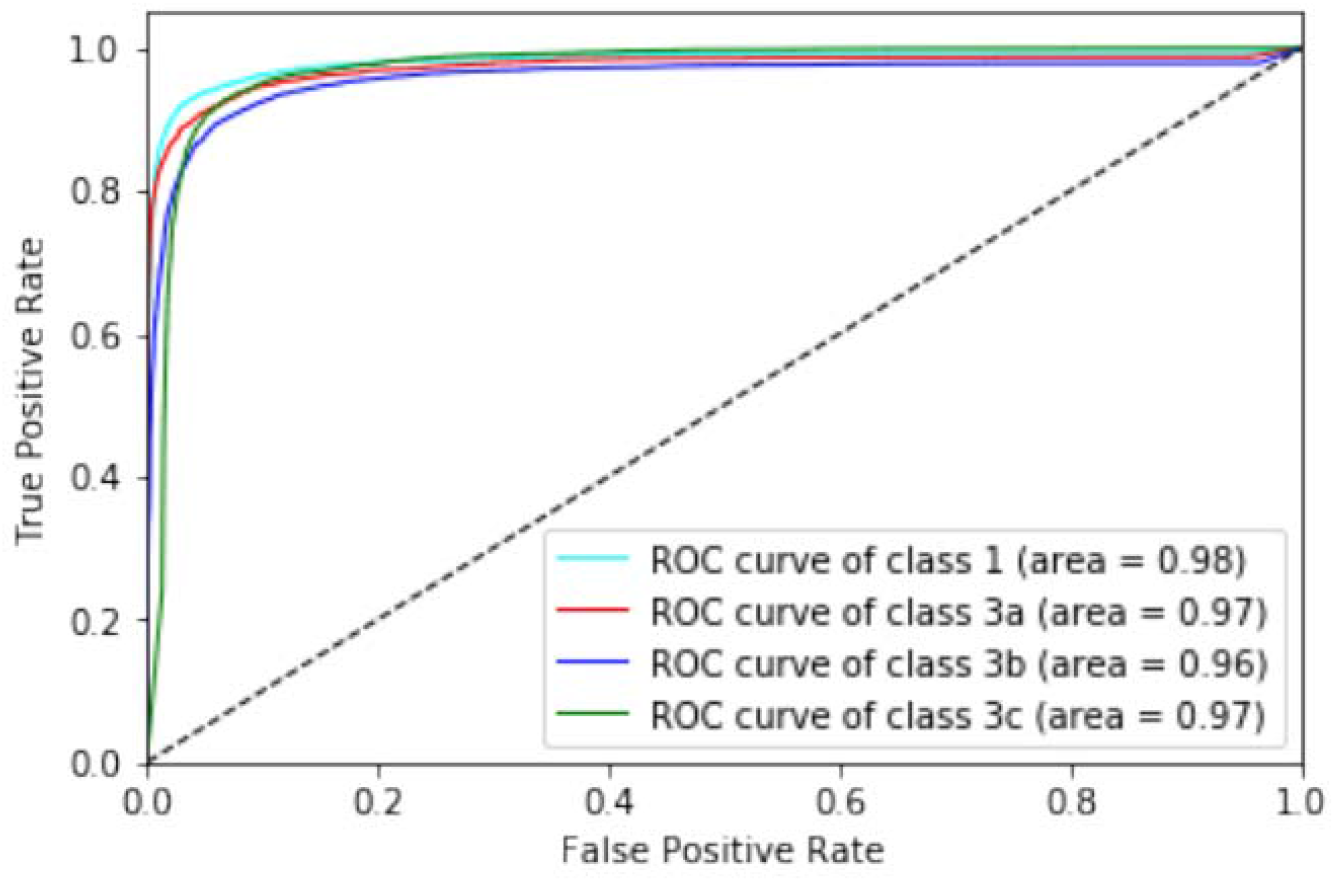
Patch-level Receiver Operating Curve (ROC) and Area Under the Curve (AUC) for different classes of celiac disease (CD) severity.

### Image Analysis Model Discerning Feature Recognition

Grad-CAMs were obtained for the patch- and WSI-level models (**Figures 4 and 5**). Notably, patch-level Grad-CAMs for Marsh IIIc found the presence of enteroendocrine cells (EECs), and on retrospective chart review, these children also had endocrine abnormalities (details in supplemental appendix **S11**). WSI Grad-CAMs focused more on the surface epithelium for Marsh score I compared to more severe scores (Marsh IIIa, IIIb, and IIIc), as shown in **Figure 5**. Detailed Grad-CAM findings are mentioned in supplemental appendix **S12**.

**Figure 4:**
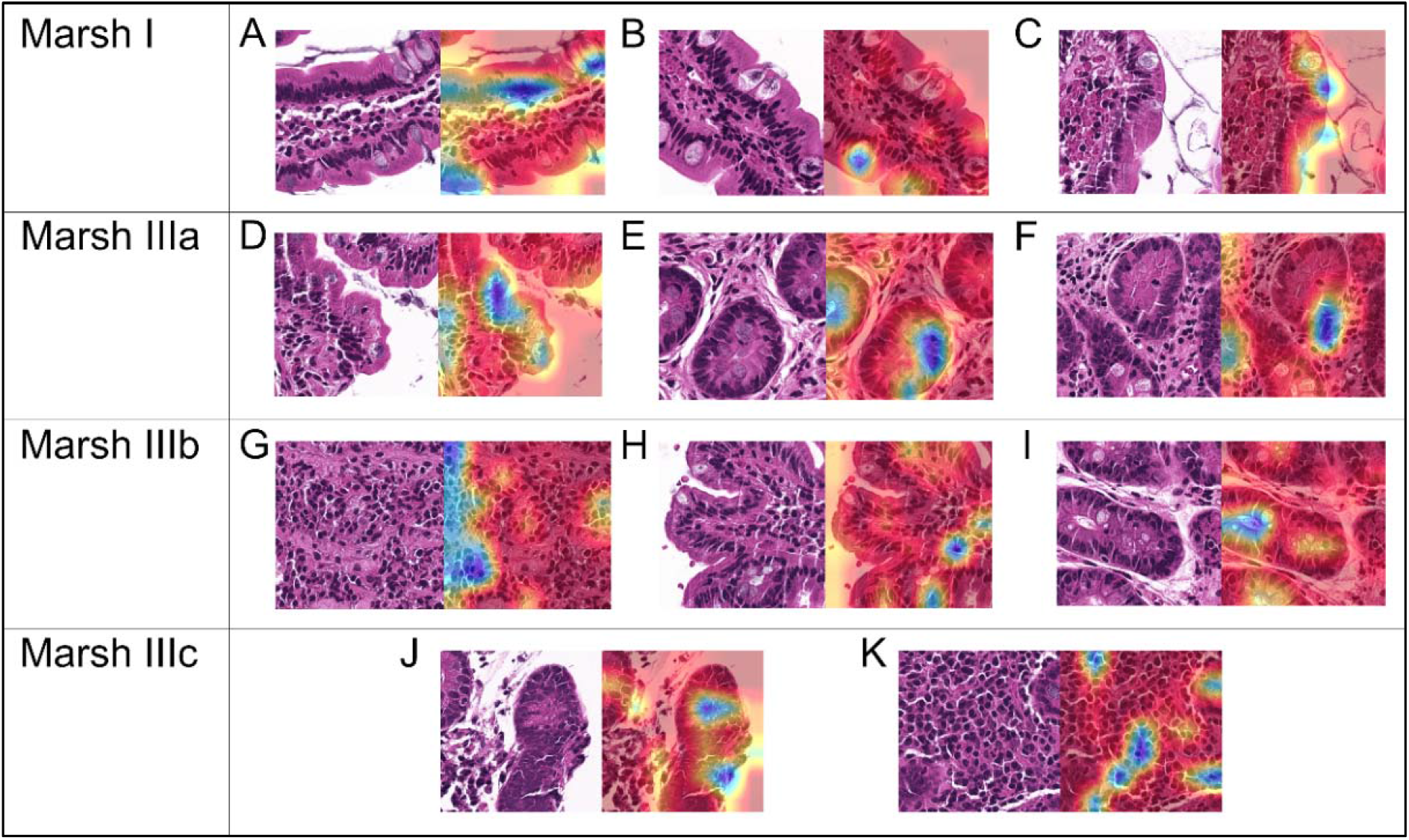
Gradient-weighted Class Activation Mapping (Grad-CAM) visualizations of the patch-level models’ output. Histologic features highlighted by the Grad-CAMs are: - <u>Marsh I:</u> (A) Tall Border Surface Epithelial Cells; (B) Goblet Cells within the Epithelium; (C) Brush Border Epithelium - <u>Marsh IIIa:</u> (D) Artefactual Separation of Tissue; (E) Cytoplasm in Crypt Epithelium; (F) Crowded Nuclei - <u>Marsh IIIb:</u> (G) Mononuclear Inflammatory Cells in Lamina Propria (LP); (H) LP between cells; (I) Central Crypt Clearing - <u>Marsh IIIc:</u> (J) Enteroendocrine Cells; (K) Granular Area (cytoplasm/connective tissue) between cells

**Figure 5:**
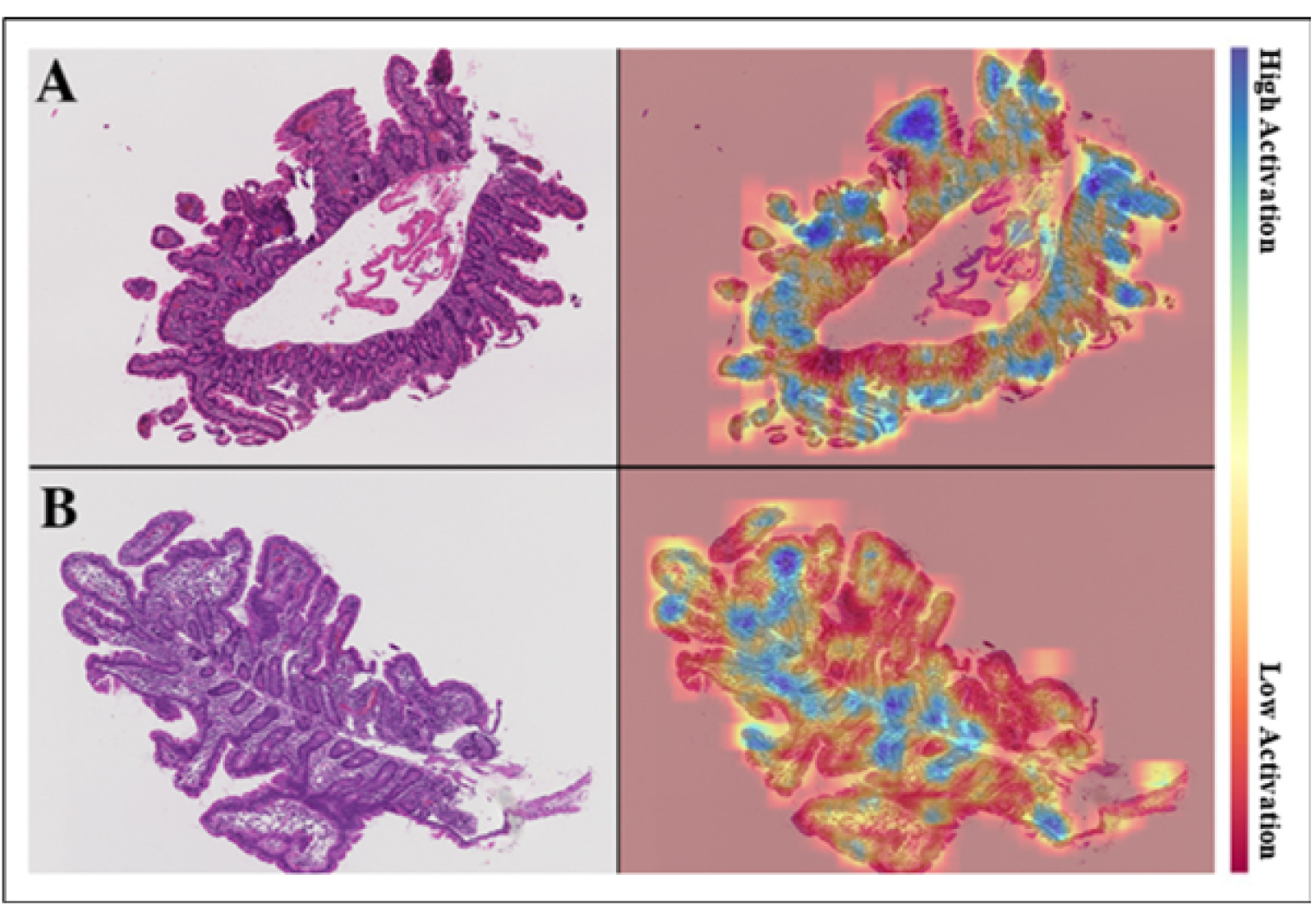
Gradient-weighted Class Activation Mapping (Grad-CAM) visualizations of the Whole Slide Image (WSI)-level models’ output.

### Immunohistochemistry (IHC) Analysis

IHC was done to confirm the EEC Grad-CAM findings in Marsh IIIc biopsies of children with endocrine abnormalities. The comparison group included children with Marsh IIIc without endocrine abnormalities. Qualitative IHC showed that chromogranin A was more localized to EECs for children with Marsh IIIc with endocrine abnormalities versus the comparison group (**Figure 6**). Quantitative assessment was performed using a Gaussian Mixture Model (GMM) to detect EECs based on clusters of stain colors per pixel. The EEC color cluster, represented as dark gray (**Figure 7**), had a higher percentage of color pixels for children with Marsh IIIc and endocrine abnormalities than those without endocrine complications (details in supplemental appendix **S13**).

**Figure 6:**
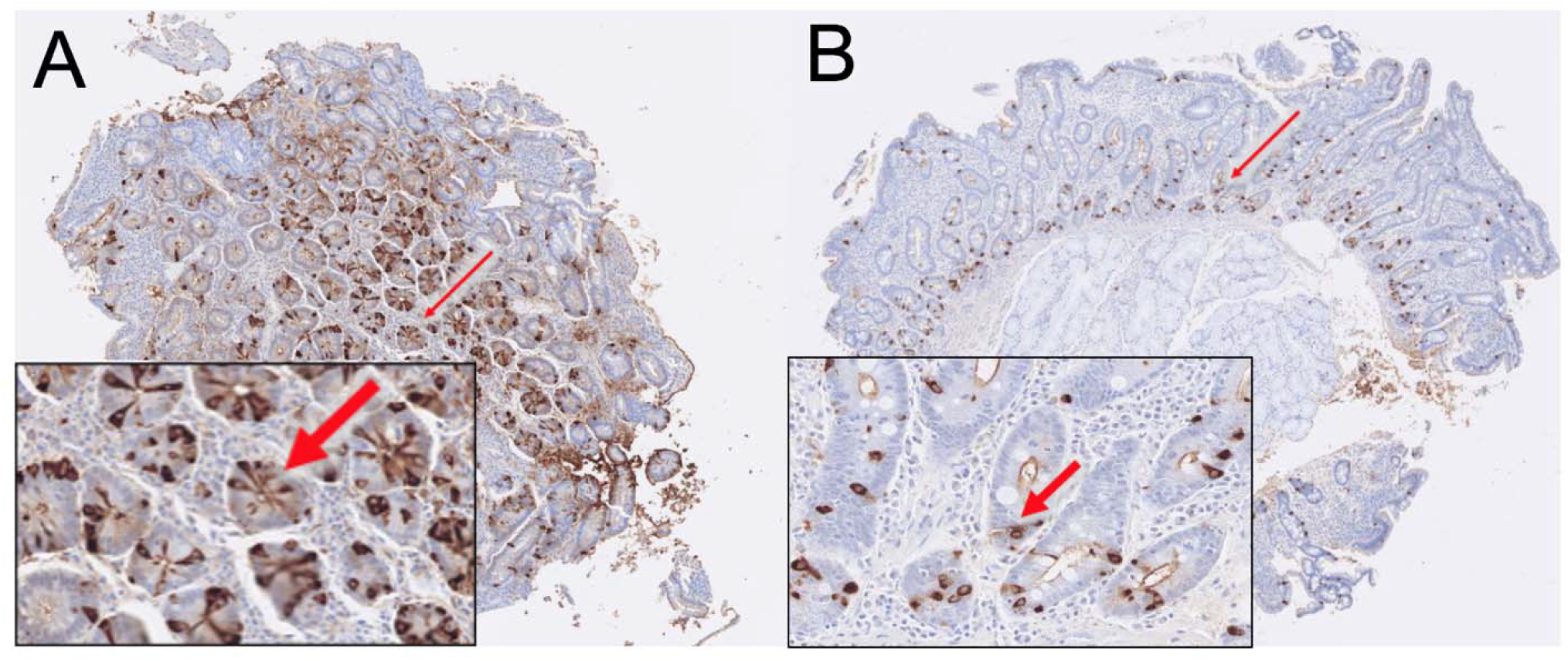
Immunohistochemistry (IHC) using chromogranin A for enteroendocrine cells (EECs) for patients with (Panel A) and without (Panel B) endocrine complications.

**Figure 7:**
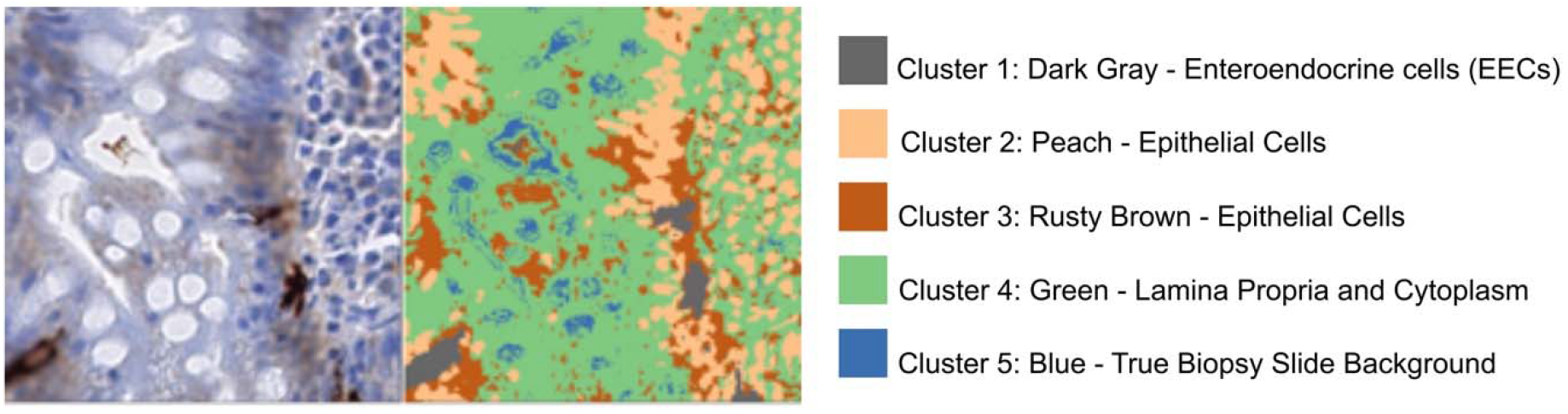
Clustering of each image pixel based on stain color using a Gaussian Mixture Model (GMM) with five clusters. Enteroendocrine cells were IHC-stained with dark brown color (on the left) which was isolated as the dark gray cluster shown in the GMM representation (on the right).

## Discussion

Once thought to be rare, Celiac Disease (CD) affects approximately 1% of the global population [24]. It appears to be associated with increased mortality along with substantial morbidity, much of which may be preventable or reversible with a gluten-free diet [add ref]. The current gold standard for CD diagnosis involves assessing proximal small intestinal biopsies [1, 2, 3] with the lack of specific histopathological markers associated with either CD severity or comorbid conditions. CD severity assessment is also challenging and falls privy to interobserver variability and non-specificity of discerning histopathological features [6, 7]. Due to this, there is a need to refine risk stratification and prediction for those who will develop severe CD or comorbidities. We utilized a deep learning-based image analytics model to assess of disease severity and identify of histopathological markers specific for certain disease severity classes and phenotype with endocrinopathies.

Our deep learning model for CD classification demonstrated accuracy comparable with a previously published study by Wei et al. [16]. This group also utilized a CNN similar to our model, although the classification was limited to CD, normal, and nonspecific duodenitis with accuracies of 95.3%, 91.0%, and 89.2%, respectively. We further categorized our CD versus normal model for CD severity classes. We also added a layer of explainability regarding the features deemed important by the model for classification with the use of Grad-CAMs.

Applying our benchmark method using Grad-CAMs [20] for discerning features specific for each severity class, we found EECs as potential biomarkers for children with severe disease (Marsh IIIc) and endocrinopathies (Type 1 diabetes and endocrine abnormalities). These markers were further confirmed via immunohistochemistry. EECs have been reported to be increased in duodenal biopsies from patients with refractory CD [25] and produce insulin or insulin-like hormones [26, 27]. They have also been linked to the development of diabetes via the Neurogenin-3 protein encoded by the NEUROG3 gene expressed in progenitor cells required for intestinal enteroendocrine cell development [28]. The proximity between enteroendocrine and immune cells in the gastrointestinal mucosa has also suggested that the interaction between the endocrine and immune systems may modulate inflammatory response in CD.

Recently European guidelines suggest that CD diagnosis can be accurately established with or without duodenal biopsies if the given recommendations are followed [11]. Our findings suggest a possible need for requirement of biopsies if not for diagnosis but for risk stratification and prediction of concurrent endocrinopathies using histopathologic markers.

Major strengths of our study include the classification of CD based on histological severity using the Marsh–Oberhüber classification. A wide variety of performance metrics (e.g., accuracy, sensitivity/specificity, PPV/NPV, F1 score) were also used to assess the deep learning model’s validity, something often missing from work done on deep learning in medicine. Moreover, our use of Grad-CAMs will assist in increasing physician confidence in deep learning-based decision making and paving the way for discerning disease-specific histopathologic markers. However, despite these strengths, we experienced limitations as well. Our study design primarily does not account for direct comparisons of human interpreters to our deep learning models, and our comparisons to human results are indirect. Additionally, there were slight variations in H&E staining chemical composition over the years within our institute, resulting in visible H&E color differences. We performed stain color normalization in an attempt to mitigate this data bias. Further, Chromogranin A also stained tissue edges along with EECs, but since this artefactual staining for equal for both comparison groups, it may not have led to the percentage of EEC color pixel-based quantification (GMM) for the group with Marsh IIIc and endocrine complications.

We have successfully developed a deep learning image analytics platform that enables visualization of discerning disease features. Using IHC, the deep feature, EEC, identified specifically for severe CD along with Type 1 diabetes and thyroid abnormalities, was confirmed via IH. We believe these findings have a broad application in the field of personalized medicine as they relate to risk stratification and patient prognostications. Additionally, it might suggest a need for biopsies at the time of diagnosis for risk and comorbidity prediction despite the presence of strong serologic markers that already confirm the presence of disease. Further work to assess whether the histopathologic markers align with gene expression signatures and external validation models are underway.

## Data Availability

Data can be available on request

## Author contributions

Concept and design: Syed, Brown, Ehsan, Khan, Sali.

Acquisition, analysis, or interpretation of data: Syed, Brown, Ehsan, Khan, Sali, Catalano, Kowsari, Cheng, Pramoonjago, Raghavan, DeBoer, Moskaluk.

Drafting of the manuscript: Syed, Ehsan, Sali.

Critical revision of the manuscript for important intellectual content: Syed, Brown, Ehsan, Khan, Raghavan, Silvester, Cheng, DeBoer, Moskaluk, Moore.

Statistical analysis: Ehsan, Sali, Adorno. Obtained funding: Syed, Brown.

Administrative, technical, or material support: Syed, Ehsan, Khan, Sali, Adorno, Pramoonjago. Supervision: Syed, Brown.

## Acknowledgments

The authors would like to acknowledge the members of Gastro Data Science Lab for their extensive feedback and continued support for this work.

## References

1 Hill ID, Dirks MH, Liptak GS, Colletti RB, Fasano A, Guandalini S, et al. Guideline for the diagnosis and treatment of celiac disease in children: recommendations of the North American Society for Pediatric Gastroenterology, Hepatology and Nutrition. Journal of pediatric gastroenterology and nutrition 2005;40:1–19.

2 Kumar V, Abbas A, Fausto N, Aster J. Robbins and Cotran Pathologic Basis of Disease, Professional. Philadelphia: Saunders Elsevier) Available from: http://books.google.com/books, 2009.

3 Mills SE, Carter D, Greenson JK, Reuter VE, Stoler MH. Sternberg’s diagnostic surgical pathology: Lippincott Williams & Wilkins, 2012.

4 Oberhuber G. Histopathology of celiac disease. Biomedicine & pharmacotherapy 2000;54:368–72.

5 Modified Marsh Classification of histologic findings in celiac disease (Oberhuber). Surgical Pathology Criteria Stanford Medicine.

6 Arguelles-Grande C, Tennyson CA, Lewis SK, Green PH, Bhagat G. Variability in small bowel histopathology reporting between different pathology practice settings: impact on the diagnosis of coeliac disease. Journal of clinical pathology 2012;65:242–7.

7 Corazza GR, Villanacci V, Zambelli C, Milione M, Luinetti O, Vindigni C, et al. Comparison of the interobserver reproducibility with different histologic criteria used in celiac disease. Clinical Gastroenterology and Hepatology 2007;5:838–43.

8 Adelman DC, Murray J, Wu T-T, Mäki M, Green PH, Kelly CP. Measuring change in small intestinal histology in patients with celiac disease. American Journal of Gastroenterology 2018;113:339–47.

9 Akirov A, Pinhas-Hamiel O. Co-occurrence of type 1 diabetes mellitus and celiac disease. World journal of diabetes 2015;6:707.

10 Cohn A, Sofia AM, Kupfer SS. Type 1 diabetes and celiac disease: clinical overlap and new insights into disease pathogenesis. Current diabetes reports 2014;14:517.

11 Al-Toma A, Volta U, Auricchio R, Castillejo G, Sanders DS, Cellier C, et al. European Society for the Study of Coeliac Disease (ESsCD) guideline for coeliac disease and other gluten-related disorders. United European gastroenterology journal 2019;7:583–613.

12 Beck AH, Sangoi AR, Leung S, Marinelli RJ, Nielsen TO, Van De Vijver MJ, et al. Systematic analysis of breast cancer morphology uncovers stromal features associated with survival. Science translational medicine 2011;3:108ra13–ra13.

13 Bejnordi BE, Veta M, Van Diest PJ, Van Ginneken B, Karssemeijer N, Litjens G, et al. Diagnostic assessment of deep learning algorithms for detection of lymph node metastases in women with breast cancer. Jama 2017;318:2199–210.

14 Doyle S, Hwang M, Shah K, Madabhushi A, Feldman M, Tomaszeweski J. Automated grading of prostate cancer using architectural and textural image features. 2007 4th IEEE International Symposium on Biomedical Imaging: From Nano to Macro: IEEE, 2007:1284–7.

15 Cruz-Roa A, Basavanhally A, González F, Gilmore H, Feldman M, Ganesan S, et al. Automatic detection of invasive ductal carcinoma in whole slide images with convolutional neural networks. Medical Imaging 2014: Digital Pathology: International Society for Optics and Photonics, 2014:904103.

16 Wei JW, Wei JW, Jackson CR, Ren B, Suriawinata AA, Hassanpour S. Automated detection of celiac disease on duodenal biopsy slides: A deep learning approach. Journal of pathology informatics 2019;10.

17 Krizhevsky A, Sutskever I, Hinton GE. Imagenet classification with deep convolutional neural networks. Advances in neural information processing systems, 2012:1097–105.

18 Simonyan K, Zisserman A. Very deep convolutional networks for large-scale image recognition. arXiv preprint arXiv:14091556 2014.

19 Russakovsky O, Deng J, Su H, Krause J, Satheesh S, Ma S, et al. Imagenet large scale visual recognition challenge. International journal of computer vision 2015;115:211–52.

20 Selvaraju RR, Cogswell M, Das A, Vedantam R, Parikh D, Batra D. Grad-cam: Visual explanations from deep networks via gradient-based localization. Proceedings of the IEEE International Conference on Computer Vision, 2017:618–26.

21 Vahadane A, Peng T, Sethi A, Albarqouni S, Wang L, Baust M, et al. Structure-preserving color normalization and sparse stain separation for histological images. IEEE transactions on medical imaging 2016;35:1962–71.

22 Sali R, Ehsan L, Kowsari K, Khan M, Moskaluk CA, Syed S, et al. Celiacnet: Celiac disease severity diagnosis on duodenal histopathological images using deep residual networks. 2019 IEEE International Conference on Bioinformatics and Biomedicine (BIBM): IEEE, 2019:962–7.

23 Pedregosa F, Varoquaux G, Gramfort A, Michel V, Thirion B, Grisel O, et al. Scikit-learn: Machine learning in Python. the Journal of machine Learning research 2011;12:2825–30.

24 Singh P, Arora A, Strand TA, Leffler DA, Catassi C, Green PH, et al. Global prevalence of celiac disease: systematic review and meta-analysis. Clinical Gastroenterology and Hepatology 2018;16:823-36. e2.

25 Di Sabatino A, Giuffrida P, Vanoli A, Luinetti O, Manca R, Biancheri P, et al. Increase in neuroendocrine cells in the duodenal mucosa of patients with refractory celiac disease. The American journal of gastroenterology 2014;109:258–69.

26 Han J, Lee H-H, Kwon H, Shin S, Yoon J-W, Jun H-S. Engineered enteroendocrine cells secrete insulin in response to glucose and reverse hyperglycemia in diabetic mice. Molecular Therapy 2007;15:1195–202.

27 Billing LJ, Larraufie P, Lewis J, Leiter A, Li J, Lam B, et al. Single cell transcriptomic profiling of large intestinal enteroendocrine cells in mice–identification of selective stimuli for insulin-like peptide-5 and glucagon-like peptide-1 co-expressing cells. Molecular metabolism 2019;29:158–69.

28 Cards G. Neurog3 Gene.

