## Supplementary Material for "Prediction of Celiac Disease Severity and Associated Endocrine Morbidities through Deep Learning-based Image Analytics"

**Affiliations:**

**Supplemental Methods**

**Appendix S1. Supplemental Methods** – Assessment of Celiac Disease Severity via Marsh–Oberhüber Classification for the Duodenum.

**Appendix S2. Supplemental Methods** – Biopsy Image Patch Creation.

**Appendix S3. Supplemental Methods** – Biopsy Patch Clustering.

**Appendix S4. Supplemental Methods** – Biopsy Stain Color Normalization.

**Appendix S5. Supplemental Methods** – Patch-level ResNet50 Model.

**Appendix S6. Supplemental Methods** – Whole Slide Image (WSI)-level Model.

**Appendix S7. Supplemental Methods** – Benchmark Visualization of Model Classification Decision-Making – Gradient-weighted Class Activation Mappings (Grad-CAMs).

**Appendix S8. Supplemental Methods** – Gaussian Mixture Model (GMM).

**Supplemental Results**

**Appendix S9. Supplemental Results** – Patch-level and Whole Slide Image-level Dataset Description.

**Appendix S10. Supplemental Results** – Deep Learning Model Performance for Celiac Disease (CD) versus histologically normal duodenal controls.

**Appendix S11. Supplemental Results** – Brief Retrospective Chart Review of Children with Endocrine Abnormalities.

**Appendix S12. Supplemental Results** – Deep Learning Image Analysis Model Region of Interest Assessment (Grad-CAM findings).

**Appendix S13. Supplemental Results** – Gaussian Mixture Model (GMM) Results for Quantitative Assessment of the Enteroendocrine Cells Stained via Chromogranin A.

**Supplemental References**

**Supplemental Methods**

**Appendix S1. Supplemental Methods** – **Assessment of Celiac Disease Severity via Marsh–Oberhüber Classification for the Duodenum.**

Marsh I has been described as normal architecture with more than 30 lymphocytes interspersed between 100 villous surface epithelial cells. Marsh II has shown to include increased intraepithelial lymphocytes (>30) along with crypt hypertrophy; although it is rarely encountered in clinical practice since patients proceed rapidly from Marsh I to IIIa. Marsh III has been further sub-divided into IIIa (partial villous atrophy), Marsh IIIb (subtotal villous atrophy), and Marsh IIIc (complete villous atrophy) [1, 2].

**Appendix S2. Supplemental Methods** – **Biopsy Image Patch Creation.**

Each WSI was divided into 500 x 500 pixel patches using a sliding window method with 50% overlap within the patches. These were further resized after data augmentation to 224 × 224 pixels for maximization of computational resources as final input for the model.

**Appendix S3. Supplemental Methods** – **Biopsy Patch Clustering.**

A two-step approach was deployed for clustering of biopsy patches such that areas with $\geq$50% were grouped together. First, a convolutional autoencoder was employed which learned the embedded features of each patch. The second step utilized a k-means clustering algorithm to cluster the embedded biopsy image features into clusters. Cluster containing patches with $\geq$50% background were not used as input for the model in order to avoid undue bias as the model might have classified based on the area of background present rather than histopathological features. Detailed computational methods have been previously published [3].

**Appendix S4. Supplemental Methods** – **Biopsy Stain Color Normalization.**

Method described by Vahadane et al. [4] preserves structural information and involves empiric selection of a target biopsy image as a reference to normalize coloration across all biopsy images. With this method, the color of all the biopsy images became the same as that of the empirically selected target biopsy image. This was an added layer of mitigating data bias as all our biopsy images were obtained from the same institute but may have varied stain chemical compositions over time.

**Appendix S5. Supplemental Methods** – **Patch-level ResNet50 Model.**

ResNet50 is a widely used deep CNN architecture with 50 layers for image classification requiring identification of microscopic patterns. We modified the final decision layers of ResNet50 to improve accuracy. To combat data sparsity we used transfer learning, an established method used to improve training using limited datasets by pre-training the model on the ImageNet dataset.

We customized the Resnet50 by removing fully connected layers and keeping only the ResNet backbone as a feature extractor. Then we added one fully connected layer with 1024 neurons that received the flattened output of the feature extractor. Finally, the output layer was added such that it represented a prediction probability for each of the four Marsh score classes: I, IIIa, IIIb, and IIIc. We used dropout on the fully-connected layers with p = 0.5 as the regulizer. The detailed computational methods of our patch-level model have been previously published [3].

**Appendix S6. Supplemental Methods** – **Whole Slide Image (WSI)-level Model.**

The ResNet50 model used was trained to classify small patches rather than WSIs. To achieve WSI-level classification, a heuristic method was developed which aggregated patch-level classifications and translated them to whole-slide inferences. Each WSI in the test set was initially patched, those patches which did not contain any information were filtered out and finally stain color normalization was performed. After these preprocessing steps our trained model was applied with the goal of image classification. For each patch, the model gives an output of a vector composed of four components showing probabilities for each one of the four classes of CD severity. The detailed computational methods of our WSI-level model have been previously published [3].

**Appendix S7. Supplemental Methods** – **Benchmark Visualization of Model Classification Decision-Making – Gradient-weighted Class Activation Mappings (Grad-CAMs).**

Gradient-weighted Class Activation Mappings (Grad-CAMs) generate the localization heatmaps highlighting specific regions of interest [5]. Activation values from an intermediate convolution layer and corresponding gradients as described by Selvaraju et al. [5] to generate these heatmaps.

**Appendix S8. Supplemental Methods** – **Gaussian Mixture Model (GMM).**

GMMs are unsupervised clustering algorithms where each cluster is modeled as a separate Gaussian distribution [6]. Cluster labels are assigned to each data point based which Gaussian has the highest likelihood that the data point is sampled from that distribution. The data points for this problem are samples of pixel colors from IHC images. The GMM was trained with the expectation-maximization algorithm to find optimal distributions for a mixture of five Gaussians.

**Supplemental Results**

**Appendix S9. Supplemental Results – Patch-level and Whole Slide Image-level Dataset Description**

For the patch-level model, patches created from WSIs of 63 patients were used as the training and test set with a 70:30 split. After application of sliding window for patching the WSIs and pre-processing the biopsy image patches (rotation, clustering, and stain color normalization), there were 28,667 biopsy image patches which fulfilled the criteria to be used in the model (<50% background). These same image patches were then aggregated (explained in supplemental methods) for the WSI-level model to provide inference at the level of an individual patients. For the four classes of Marsh I, IIIa, IIIb, and IIIc, there were 20, 21, 44, and 35 WSIs, respectively.

**Appendix S10. Supplemental Results – Deep Learning Model Performance for Celiac Disease (CD) versus histologically normal duodenal controls.**

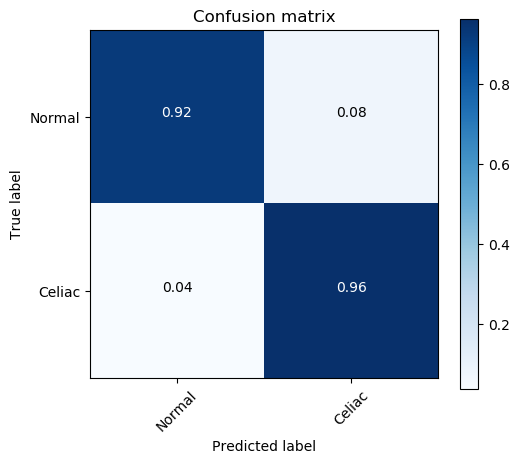
The model for CD versus histologically normal controls demonstrated an accuracy of 94% with confusion matrix for the performance accuracy shown below.

**Appendix S11. Supplemental Results** – **Brief Retrospective Chart Review of Children with Endocrine Abnormalities.**

| **Child** | **Endocrine Abnormalities and other clinical characteristics** |
| --- | --- |
| 1 | Type 1 Diabetes, Hypothyroidism |
| 2 | High risk for diabetes and hypothyroidism due to HLA makeup, two Thyroid Stimulating Hormone labs with low and normal values and normal Free T4 labs |
| 3 | Trisomy 21, Hypothyroidism |
| Brief retrospective chart review of children with endocrine abnormalities along with their other clinical characteristics. | |

**Appendix S12. Supplemental Results** – **Deep Learning Image Analysis Model Region of Interest Assessment (Grad-CAM findings).**

324 Grad-CAMs were reviewed (5-6 patches per WSI) and they focused on medically relevant features for each Marsh score class. The model identified brush border epithelium primarily on tall surface epithelial cells and goblet cells within the surface and crypt epithelium for Marsh I. For Marsh IIIa, the model visualized artefactual separation of tissue and pink dense cytoplasm within the crypt epithelium. For Marsh IIIb, defining features included pink dense cytoplasm within the crypt epithelium similar to Marsh IIIa, as well as goblet cells within the crypt epithelium and crypt epithelium only (as opposed to Marsh I which also focused on villous surface epithelium and goblet cells within the surface epithelium). For Marsh IIIc, central granular areas between cells were highlighted. Notably, for Marsh IIIc the model focused on all the areas within Grad-CAMs that contained enteroendocrine cells. For the WSI-level model, Grad-CAMs focused more on the surface epithelium for Marsh score I compared to more severe scores (Marsh IIIa, IIIb, and IIIc).

**Appendix S13. Supplemental Results – Gaussian Mixture Model (GMM) Results for Quantitative Assessment of the Enteroendocrine Cells Stained via Chromogranin A.**

Quantitative assessment was performed by detecting EECs in each image pixel with Gaussian Mixture Model (GMM). The GMM identified five major clusters that represent differences in biopsy features based solely on the stain color of the IHC-stained biopsy images. The EEC color cluster was easily isolated because of the dark brown IHC stain appearance and was represented as dark gray color on the clustering example image in the main manuscript. This color cluster had a higher percentage of color pixels for children with Marsh IIIc and endocrine abnormalities than those without endocrine complications (see table below).

| **Disease Status** | **Percentage* of Enteroendocrine Cells** |
| --- | --- |
| With endocrine complications | 10.3% |
| Without endocrine complications | 7.3% |
| *Percentage calculated via the following equation: (sum of pixels of cluster 1)/(sum of pixels of clusters 1, 2, 3, and 4)  Cluster 1: Dark Gray – Enteroendocrine cells (EECs)  Cluster 2: Peach – Epithelial Cells  Cluster 3: Rusty Brown – Epithelial Cells Cluster 4: Green – Lamina propria and Cytoplasm  Cluster 5: Blue – Biopsy Slide Background | |

**Supplemental References**

1	Modified Marsh Classification of histologic findings in celiac disease (Oberhuber). Surgical Pathology Criteria Stanford Medicine.

2	Oberhuber G. Histopathology of celiac disease. Biomedicine & pharmacotherapy 2000;**54**:368-72.

3	Sali R, Ehsan L, Kowsari K, Khan M, Moskaluk CA, Syed S*, et al.* Celiacnet: Celiac disease severity diagnosis on duodenal histopathological images using deep residual networks. 2019 IEEE International Conference on Bioinformatics and Biomedicine (BIBM): IEEE, 2019:962-7.

4	Vahadane A, Peng T, Sethi A, Albarqouni S, Wang L, Baust M*, et al.* Structure-preserving color normalization and sparse stain separation for histological images. IEEE transactions on medical imaging 2016;**35**:1962-71.

5	Selvaraju RR, Cogswell M, Das A, Vedantam R, Parikh D, Batra D. Grad-cam: Visual explanations from deep networks via gradient-based localization. Proceedings of the IEEE International Conference on Computer Vision, 2017:618-26.

6	Pedregosa F, Varoquaux G, Gramfort A, Michel V, Thirion B, Grisel O*, et al.* Scikit-learn: Machine learning in Python. the Journal of machine Learning research 2011;**12**:2825-30.
